# Determinants of Health Literacy and their Impact on Residents in Newham, London

**DOI:** 10.1101/2025.08.26.25334435

**Authors:** Nathan Green, Ysabella-Rozetta Hawkings

## Abstract

This study investigates the determinants of health literacy in Newham, London, using a statistical modelling approach. Health literacy is defined using literacy, numeracy, and Information and Communication Technology (ICT) literacy from the ONS Skills for Life (SfL) survey 2011. The research combines survey, the Newham Residents Survey (NRS), and the UK 2021 Census data. Multilevel regression with poststratification (MRP) and average treatment effects (ATE) are used to quantify the impact of various factors on health literacy at the local level.

The study identifies several significant determinants of health literacy, including age, ethnicity, qualification level, English as a first language, job status, gross income, and home ownership. Specifically, individuals aged 45 years and over are estimated to have lower ICT health literacy. White ethnicity is associated with higher numeracy scores. Additionally, health literacy worsens as area deprivation increases. Qualification level is estimated to be the most influential factor across all health literacy outcomes. The ultimate aim of this study is to inform targeted health literacy interventions at the local level by quantifying the impact of key determinants with uncertainty, thereby aiding in the prioritisation of resources.

## Introduction

Health literacy is broadly defined as the ability to access, understand, appraise, and communicate health information, enabling individuals to engage in healthcare and maintain good health throughout their lives. Low health literacy is a significant concern and has received greater attention in recent years. It is associated with difficulties in making informed healthcare decisions, leading to poorer access to, knowledge, and quality of care.

Traditionally, health literacy was often assessed using unidimensional measures focused primarily on generic literacy and numeracy skills. However, there is a growing recognition of the multidimensional nature of health literacy, encompassing factors such as social support, navigation of healthcare systems, and communication skills. This broader perspective, as captured by tools like the influential Health Literacy Questionnaire (HLQ) Osborne et al (2013), emphasizes the complex interplay of factors that influence an individual’s capacity to manage their health effectively.

Broadly, disparities in health outcomes have been linked to wider *social determinants of health (SDoH)*, which are non-medical social, environmental and economic factors that have been shown to impact up to 90% of health status and disparities Woolf (2019). Health literacy itself is intricately related to SDoH, conceptualised in the literature in various ways: most commonly as a result of SDoH, but also as a mediator between other SDoH and health outcomes, or even as a modifiable in its own right Stormacq et al (2019).

Previous research has identified numerous socio-demographic factors consistently associated with lower levels of health literacy, such as lower educational attainment, older age, lower income, and racial/ethnic minority status Dolezel and Hewitt (2023); Martin et al (2009); Stormacq et al (2019). Education, in particular, is frequently cited as the most important determinant of health literacy Stormacq et al (2019); Laursen and Seed (2016). Other associated factors include gender, marital status, depression, chronic conditions, and area deprivation Lee et al (2015); Sun et al (2022); Dolezel and Hewitt (2023); Martin et al (2009); Stormacq et al (2019). Thinking has also shifted in recent years from it being wholly dependent on the individual to now recognise the role of the system and institutions in improving health literacy.

Research has shown a strong link between lower health literacy and poorer health outcomes, with studies using literacy and numeracy as proxies. A meta-analysis by Zhang et al (2014) found that health literacy influences physical health, treatment adherence, and health behaviours, with lower levels linked to higher hospitalization rates, poor aftercare engagement, and negative attitudes toward seeking treatment.

There are several challenges and potential stigma associated with directly measuring health literacy in large populations or clinical settings, along with the time required Laursen and Seed (2016); Martin et al (2009). To address this, predictive models utilising available socio-demographic data have been explored to estimate health literacy levels and identify communities or individuals at higher risk Martin et al (2009); Laursen and Seed (2016); Campbell et al (2019). Building on this previous work, this paper develops methods applied to social determinants of health and health literacy. We will adopt a statistical modelling approach with methods borrowed from causal inference and economics, and drawing on data from multiple sources.

This study focusses on Newham, a diverse borough in East London that faces unique challenges. Newham has been identified as having some of the lowest levels of health literacy in the UK University of Southampton and NHS England (n.d.); Laursen and Seed (2016), a situation potentially compounded by factors such as a high prevalence of residents who speak English as a second language, its ranking as one of the most deprived boroughs in London, and the complexities of multigenerational or multiple-occupancy households. To address this urgent need for better understanding, this study investigates the key determinants of health literacy in Newham. These findings can be used to produce rankings of the determinants in terms of their potential impact. Intervention prioritisation can target subpopulations to make best use of available resources and funds. The aim of this work is to investigate health literacy in Newham as an exemplar of other local areas incorporating associated uncertainties, to understand the relative importance of population characteristics, and inform which health literacy interventions may be most appropriate and impactful for a local population.

## Methods

### Data

There are two types of data sets used in this study; Individual-level survey data to model associations between covariates and health literacy, and local-level data for the distribution of covariates in Newham. The latter comes from several different sources.

#### ONS Skills for Life (SfL) 2011 survey

The ONS Skills for Life (SfL) 2011 survey BIS Research Paper Number 81 (2012) was a comprehensive computer-based assessment conducted by the ONS to evaluate literacy, numeracy, and ICT (information and communication technology) skills of adults in England. The survey aimed to provide insights into the proficiency levels of adults aged 16-65 years old and to inform policy decisions and educational programs aimed at improving these essential skills.

The survey sampled 7230 participants in total. Multiple covariates were collected as part of the SfL survey. The survey data used in this work follows the analysis performed in Rowlands et al (2015) and we use the same set of covariates identified as significant: age in years (16-44, ≥45), sex (male, female), ethnicity (white, non-white), job status (routine/manual/students/unemployed, intermediate, managerial/professional), working status (yes, no), UK born (yes, no), home ownership (yes, no), English as first language (yes, no), qualification level (5 grade A–C GCSE or above, below 5 grade A–C GCSE), gross annual income (<£10,000, ≥£10,000) and Index of Multiple Deprivation (IMD). The survey provides raw IMD scores and groups according to the distributions of health literacy outcomes. We grouped the IMD data by quintiles to allow separation in the data because some of the group frequencies were very small. Other data collected that were not used included household composition, region of residence, occupation and industry sector, socioeconomic classification, self-reported health status, disability status.

#### Literacy, numeracy and ICT outcomes

The SfL survey tested the ability to read and understand text, calculate and manipulate numbers, extract and interpret information, and understand a range of vocabulary. They were also tasked with using a word processor, composing and sending emails, and using spreadsheets. Participants were graded against the five lowest levels of the National Qualifications Framework (NQF) UK Government (2024). The NQF levels are EL1 and EL2 (Entry Level 1 and 2): These are a basic literacy level; EL3 (Entry Level 3): individuals at this level can understand and use simple texts; L1 (Level 1): This is considered the foundational level of literacy; L2 or above (Level 2 or above): This indicates a higher level of literacy proficiency. Individuals at Level 2 can deal with a wide range of written texts.

#### Newham Residents Survey (NRS)

The Newham Residents Survey (NRS) Opinion Research Services (2023) is a periodic survey, usually every two years, conducted to gather detailed information on the views, experiences, and needs of residents in the London Borough of Newham. The survey covers various aspects of living in Newham, including satisfaction with local services, community safety, health and well-being, housing, and employment. This helps the local authority to make informed decisions and tailor services to better meet the needs of the community. Up until 2019, the survey was performed face to face, but due to COVID-19 in 2021 this switched to address-based online surveying, and the same survey method was used in 2023 due to cost and survey comparability. This means that 2021 is the baseline to monitor change. Random probability sampling is used at the community neighbourhood area level. Data are weighted using UK Census (2021) and Annual Population Survey (APS) data Office for National Statistics, Social Survey Division (January 2022 December 2023) to ensure representativeness; this includes age, gender, tenure, ethnic group, and working status. The APS is a major survey series, which aims to provide data that can produce reliable estimates at local authority level. Key topics covered in the survey include education, employment, health and ethnicity. Access to the individual-level data from the NRS allows us to directly estimate the joint distribution of covariates for Newham residents. We took age, sex, ethnicity, working status and home ownership from the NRS. The survey did not directly ask for gross individual annual income. Further, the only question about English language was about proficiency and so whether an individual had English as a first language could not be determined. As a result, we could not rely solely on using the NRS and needed to use other resources.

#### Additional data sources

For IMD data, the local income deprivation data set for mid 2021 population estimates for all 164 Lower Super Output Areas (LSOAs) in Newham was obtained from the ONS website Office for National Statistics (2022). This was then aggregated for each IMD decile population sum totals.

The Labour Force Survey (LFS) is a major household survey conducted by the ONS to gather essential statistics on employment, unemployment, and economic inactivity Office for National Statistics (2024). We used Quarterly Labour Force Survey microdata from July to September 2024 to estimate the joint distribution of the variables not in the NRS (highest level of qualification, English as a first language, UK born, job status and gross income). This was calculated using a type of iterative proportional fitting (IPF) (or *raking* in survey statistics) which is a method to match the marginal distributions (i.e. Newham) with the marginals from the microdata Deming and Stephan (1940).

### Health literacy definition

Health literacy has several specific definitions Sørensen et al (2012). We will adopt the definition used in Laursen and Seed (2016) and Rowlands et al (2015), where health literacy is defined using the SfL survey variable for i) ICT skills (computer and internet use); ii) English literacy level (reading, writing, speaking and listening); iii) numeracy level (understanding and manipulating numbers).

In Rowlands et al (2015) a sample of health materials, including medicine labels, booklets, application forms and others were used which covered themes of health promotion, managing illness, systems navigation and disease prevention. All the materials were assessed for their literacy and numeracy complexity by education experts external. Education reviewers assessed the level of skill required to understand and use the materials. These were graded up to and including level 2, the level expected to be achieved by age 16 years; materials above this level were grouped with level 2.

SfL responses were mapped to the binary health literacy scale according to whether they are above or below the threshold determined by the education experts. The thresholds used were L2 for literacy, L1 for numeracy and EL3 for ICT.

Because SfL survey has previously been linked to health literacy by Rowlands et al (2015) it was natural to build on this. However, we recognise that this data set is from 2011. We make the assumption that the conditional relationship between the covariates and the health literacy outcomes has not significantly changed over time. An alternative annually collected source which contains information on literacy and numeracy in the UK is the Understanding Society UK Household Longitudinal Study. In particular, PIAAC (OECD’s Programme for International Assessment of Adult Competencies) evaluates literacy and numeracy skills in adults for many countries globally and has data for 2011 and 2022.

#### Comparison of data sets

Table 1 shows the population counts and proportions for the SfL 2011 data set and Newham Residents Survey 2023 by variable and category. The sample sizes were 5818 for literacy assessment, 4396 for numeracy assessment, and 2274 for ICT assessments. The total sample of the NRS 2023 consisted of 2270 valid records. The population distribution is similar between health literacy outcomes but markedly different for the NRS. For example, UK born for SfL is 88% or 87% whereas for Newham this is 46%. This clearly corresponds to English as first language and White ethnicity being lower for Newham at 65% (91-2% for SfL), and 30% (88-90% for SfL), respectively. IMD for Newham is concentrated in deciles 2, 3 and 4 with only a few or no counts in the others. Figure 1 shows dumbbell plots of the differences between SfL 2011 data set and Newham Residents Survey 2023 for the literacy outcome. Other outcome plots are given in the appendix.

**Table 1.**
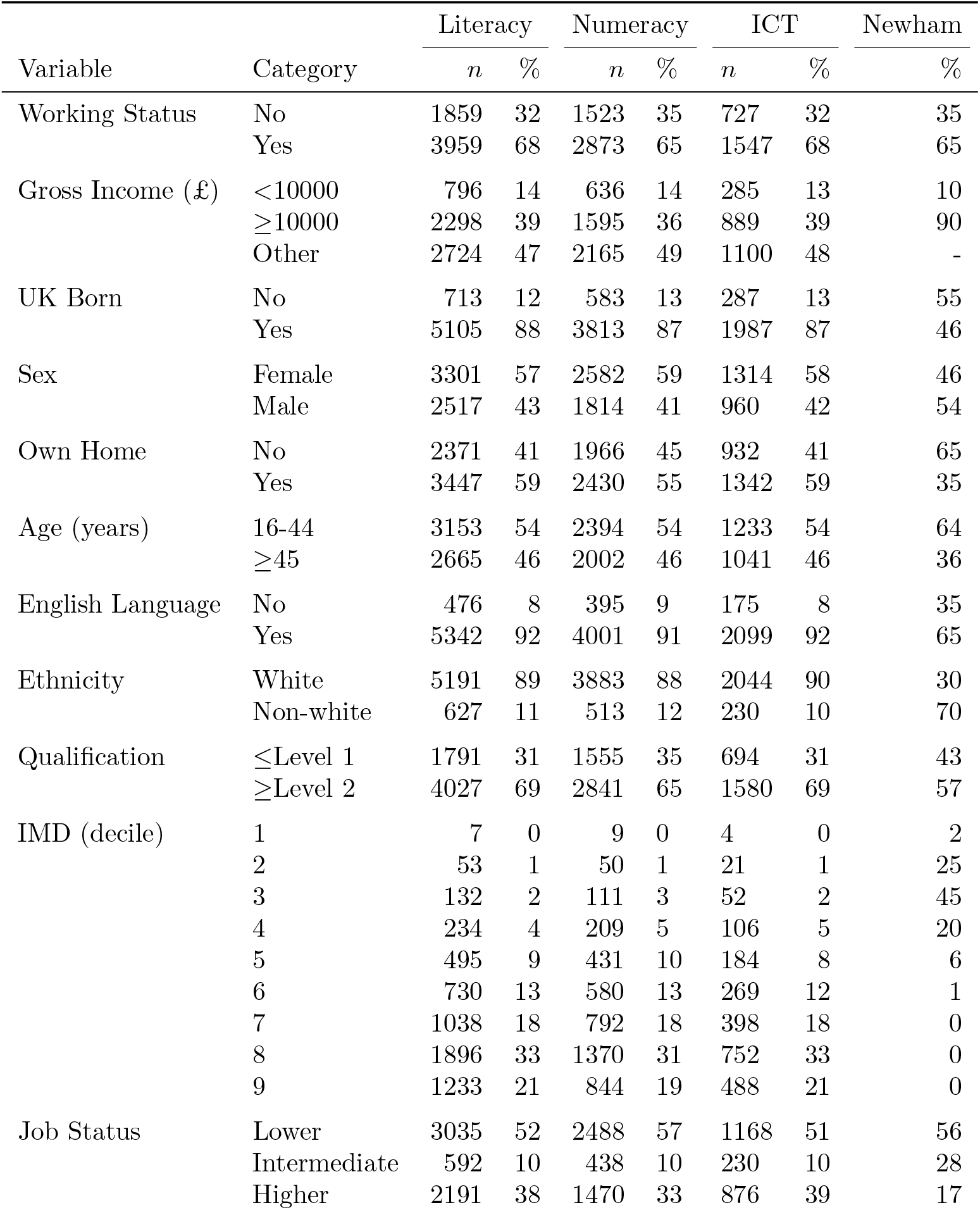
Demographic and health literacy data summary table. Literacy, Numeracy and ICT are taken from the Skills for Life Survey 2011. The Newham population proportions are taken from the Newham Residents Survey 2023, unless otherwise stated. ONS Labour Force Survey 2024 data are used for English Language, Job Status and Gross Income. The ONS local income deprivation data set for mid-2021 was used for IMD.

| Variable | Category | Literacy |  | Numeracy |  | ICT |  | Newham |
| --- | --- | --- | --- | --- | --- | --- | --- | --- |
|  |  | *n* | % | *n* | % | *n* | % | % |
| Working Status | No | 1859 | 32 | 1523 | 35 | 727 | 32 | 35 |
|  | Yes | 3959 | 68 | 2873 | 65 | 1547 | 68 | 65 |
| Gross Income (£) | <10000 | 796 | 14 | 636 | 14 | 285 | 13 | 10 |
|  | ≥10000 | 2298 | 39 | 1595 | 36 | 889 | 39 | 90 |
|  | Other | 2724 | 47 | 2165 | 49 | 1100 | 48 | - |
| UK Born | No | 713 | 12 | 583 | 13 | 287 | 13 | 55 |
|  | Yes | 5105 | 88 | 3813 | 87 | 1987 | 87 | 46 |
| Sex | Female | 3301 | 57 | 2582 | 59 | 1314 | 58 | 46 |
|  | Male | 2517 | 43 | 1814 | 41 | 960 | 42 | 54 |
| Own Home | No | 2371 | 41 | 1966 | 45 | 932 | 41 | 65 |
|  | Yes | 3447 | 59 | 2430 | 55 | 1342 | 59 | 35 |
| Age (years) | 16-44 | 3153 | 54 | 2394 | 54 | 1233 | 54 | 64 |
|  | ≥45 | 2665 | 46 | 2002 | 46 | 1041 | 46 | 36 |
| English Language | No | 476 | 8 | 395 | 9 | 175 | 8 | 35 |
|  | Yes | 5342 | 92 | 4001 | 91 | 2099 | 92 | 65 |
| Ethnicity | White | 5191 | 89 | 3883 | 88 | 2044 | 90 | 30 |
|  | Non-white | 627 | 11 | 513 | 12 | 230 | 10 | 70 |
| Qualification | ≤Level 1 | 1791 | 31 | 1555 | 35 | 694 | 31 | 43 |
|  | ≥Level 2 | 4027 | 69 | 2841 | 65 | 1580 | 69 | 57 |
| IMD (decile) | 1 | 7 | 0 | 9 | 0 | 4 | 0 | 2 |
|  | 2 | 53 | 1 | 50 | 1 | 21 | 1 | 25 |
|  | 3 | 132 | 2 | 111 | 3 | 52 | 2 | 45 |
|  | 4 | 234 | 4 | 209 | 5 | 106 | 5 | 20 |
|  | 5 | 495 | 9 | 431 | 10 | 184 | 8 | 6 |
|  | 6 | 730 | 13 | 580 | 13 | 269 | 12 | 1 |
|  | 7 | 1038 | 18 | 792 | 18 | 398 | 18 | 0 |
|  | 8 | 1896 | 33 | 1370 | 31 | 752 | 33 | 0 |
|  | 9 | 1233 | 21 | 844 | 19 | 488 | 21 | 0 |
| Job Status | Lower | 3035 | 52 | 2488 | 57 | 1168 | 51 | 56 |
|  | Intermediate | 592 | 10 | 438 | 10 | 230 | 10 | 28 |
|  | Higher | 2191 | 38 | 1470 | 33 | 876 | 39 | 17 |

**Fig. 1.**
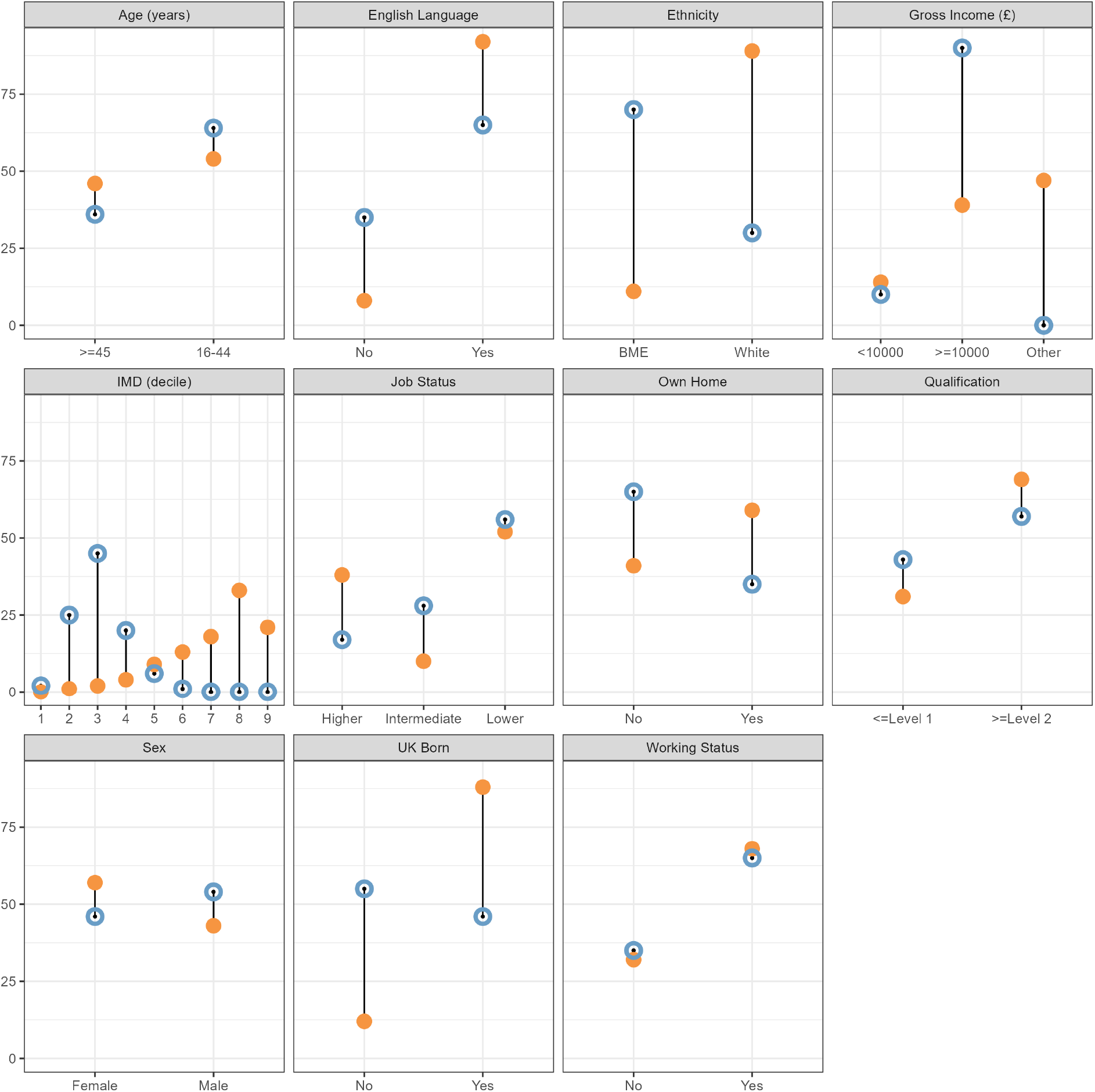
Dumbbell plots of proportions for the Skills for Life survey 2011 (solid orange) and Newham Residents Survey 2023 (hollow blue) by variable and category.

### Statistical analysis

Relevant information is available from different datasets, but each has limitations for our research question. The NRS is current and specific to Newham but lacks health literacy data. The SfL survey, conversely, provides literacy and numeracy measures that can be transformed into health literacy outcomes using Rowland *et al*. Rowlands et al (2015). The SfL survey is not specific to Newham but is from a subset of a different population. We address these limitations by combining these datasets using *Multilevel Regression with Post-Stratification (MRP)*.

#### Multilevel Regression with Post-Stratification (MRP)

MRP is a statistical technique that combines multilevel modelling with post-stratification to estimate subnational characteristics from survey data Park et al (2004, 2006). It accounts for hierarchical data structures (e.g. individuals within groups) and adjusts for known population characteristics, yielding more precise small-area estimates. MRP allows inference on a population of interest from sparse or non-representative samples, integrating small-area estimation and population adjustment techniques. While previously used in political science Lax and Phillips (2009); Ghitza and Gelman (2013), its public health applications are relatively few. MRP proceeds in two stages.

##### Multilevel Regression

A multilevel model of individual survey responses (from SfL data) is estimated to generate an estimate for each demographic group. Our model uses a binary outcome *y*_*i*_ for individual *i* (above/below health literacy threshold). The predicted probability 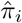 is defined as:

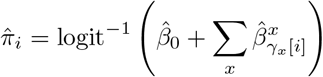

where 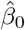 is the intercept, 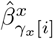 are coefficients for covariates *x* (age, sex, eng, white, ukborn, qual, inc, job, work, home), and *γ*_*x*_[*i*] represents the level or category for covariate *x* for individual *i*. IMD is included as multilevel random effects 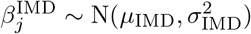. Priors distributions for fixed effects are normal distributions centered at zero with modest variance, and half-normal priors are used for random effect standard deviations Gelman (2006).

##### Post-Stratification

The health literacy probabilities for each demographic category (cell *c*) are weighted by their proportion in the actual Newham population. With 11 covariates resulting in |*S*| = 13,824 cells, the post-stratified estimate 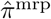 is:

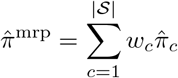

where 𝒮 is the set of all covariate combinations, *N*_*c*_ is the population frequency for cell *c, N* is the total population size, and *w*_*c*_ = *N*_*c*_*/N* are the combination weights.

#### Predictive comparisons and MRP

The *Average Marginal Effect (AME)* is commonly used to interpret regression results on the outcome scale, measuring the change in outcome probability due to an incremental change in a predictor Onukwugha et al (2015a,b); Norton et al (2019). We adopt this idea but from an Average Treatment Effect (ATE) perspective, since they can be thought of as equivalent for a binary valued treatment covariate. This approach is related to causal estimation but we adopt the term *predictive comparison* from Gelman and Pardoe (2007) to emphasize that we are summarizing the structure of the predictive model and not necessarily estimating causal effects.

We then use MRP to estimate target population specific effects with an ATE model, which we shall call *MRP-ATE*. This is an area explored by Gao Gao et al (2021). Under the potential outcome framework Hernán and Robins (2020), the ATE on the population level is 𝔼 [*Y* (1) *− Y* (0)], which averages the effect of an intervention (e.g., changing a covariate from 0 to 1) across the entire population. We define the MRP-ATE for a particular target population (different from the sample population) as

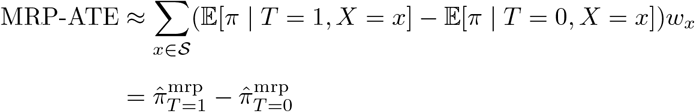

This allows us to investigate the impact of changing a covariate’s value (our “treatment”) on health literacy, even for non-modifiable factors like age or sex. For example, we can model the effect of an intervention that makes older individuals’ ICT health literacy “as-if” they were younger. We assume that the chosen covariates are appropriate confounders across all focal covariates.

#### Priority ranking

For policy applications such as healthcare cost-effectiveness, it’s useful to rank different options. We work with the *cumulative rank probability* 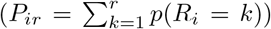, which indicates the total probability of a variable achieving a given rank or higher. A key feature of our analysis is using the *absolute treatment effect*, |*Y* (1) *− Y* (0)|, for ranking, acknowledging that the direction of the effect can be arbitrary in model choice. To summarize these probabilistic rankings, we adopt the *Surface Under the Cumulative Ranking Curve (SUCRA)* metric, common in multiple-treatment meta-analysis Salanti et al (2011). SUCRA represents the percentage of the maximum possible cumulative rank an intervention (in our case, an input variable) can achieve, providing a single value where a higher SUCRA indicates a better overall rank relative to others. For our model, it is given by the following

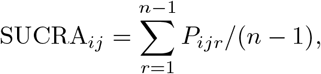

where *P*_*ijr*_ is the cumulative probability for variable *i* at level *j* and rank *r*. The mean rank is

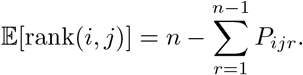

#### Software

All data preparation and analyses were performed with the statistical programming language R v.4.5.0 R Core Team (2023) calling Stan for Bayesian analyses. Code is available on GitHub at https://github.com/n8thangreen/healthliteracy. The MRP code is adapted from the code provided in Damonte and Negri (2023). IPF was performed with the *simPop* package in R Templ et al (2017).

## Results

Figure 2 shows a forest plot of MRP-ATE for the probability of being not health literate for each of the health literacy outcomes. The labels on the *y*-axis indicate the particular covariate level to which all individuals are assigned whilst averaging over all of the other covariates with Newham’s population profile. Negative values demonstrate an improvement in health literacy and positive values a worsening. The corresponding values are given in Table 2.

**Table 2.** Multilevel regression with poststratification average treatment effects (MRP-ATE) for each variable by probability not health literate outcomes ICT, literacy and numeracy. Mean change in probability of not being health literate is given, with upper and lower bounds of the 95% intervals.

| Variable/Category | Literacy | Numeracy | ICT |
| --- | --- | --- | --- |
| Age (years) |  |  |  |
| ≥45 | 0.082 [0.064, 0.103] | 0.027 [0.010, 0.050] | 0.212 [0.152, 0.267] |
| English Language |  |  |  |
| Yes | -0.186 [-0.246, -0.147] | 0.032 [0.000, 0.071] | 0.008 [-0.057, 0.062] |
| Ethnicity |  |  |  |
| White | -0.069 [-0.101, -0.030] | -0.128 [-0.172, -0.093] | -0.041 [-0.101, -0.002] |
| Gross Income (£) |  |  |  |
| ≥10000 | -0.013 [-0.036, 0.010] | -0.025 [-0.043, -0.003] | -0.055 [-0.123, -0.001] |
| Other | 0.070 [0.038, 0.102] | 0.030 [0.016, 0.051] | 0.043 [-0.005, 0.108] |
| IMD (quintile) |  |  |  |
| 2 | 0.013 [-0.116, 0.136] | 0.010 [-0.105, 0.133] | 0.021 [-0.035, 0.118] |
| 3 | 0.076 [-0.013, 0.204] | -0.001 [-0.122, 0.090] | 0.001 [-0.072, 0.075] |
| 4 | 0.012 [-0.090, 0.142] | -0.028 [-0.138, 0.053] | -0.015 [-0.093, 0.036] |
| 5 | -0.056 [-0.164, 0.075] | -0.108 [-0.213, -0.015] | -0.034 [-0.109, 0.017] |
| Job Status |  |  |  |
| Intermediate | 0.041 [0.002, 0.081] | 0.050 [0.012, 0.094] | -0.003 [-0.054, 0.041] |
| Lower | 0.155 [0.132, 0.189] | 0.124 [0.090, 0.159] | 0.113 [0.080, 0.145] |
| Own home |  |  |  |
| Yes | -0.072 [-0.090, -0.056] | -0.067 [-0.087, -0.048] | -0.024 [-0.049, 0.010] |
| Qualification |  |  |  |
| ≥level 2 | -0.224 [-0.245, -0.190] | -0.137 [-0.172, -0.106] | -0.141 [-0.180, -0.099] |
| Sex |  |  |  |
| Male | 0.036 [0.015, 0.049] | -0.083 [-0.103, -0.065] | 0.023 [-0.011, 0.046] |
| UK Born |  |  |  |
| Yes | -0.018 [-0.056, 0.034] | -0.002 [-0.029, 0.025] | -0.069 [-0.151, -0.006] |
| Working Status |  |  |  |
| Yes | 0.034 [-0.004, 0.063] | 0.002 [-0.029, 0.029] | -0.012 [-0.050, 0.017] |

**Fig. 2.**
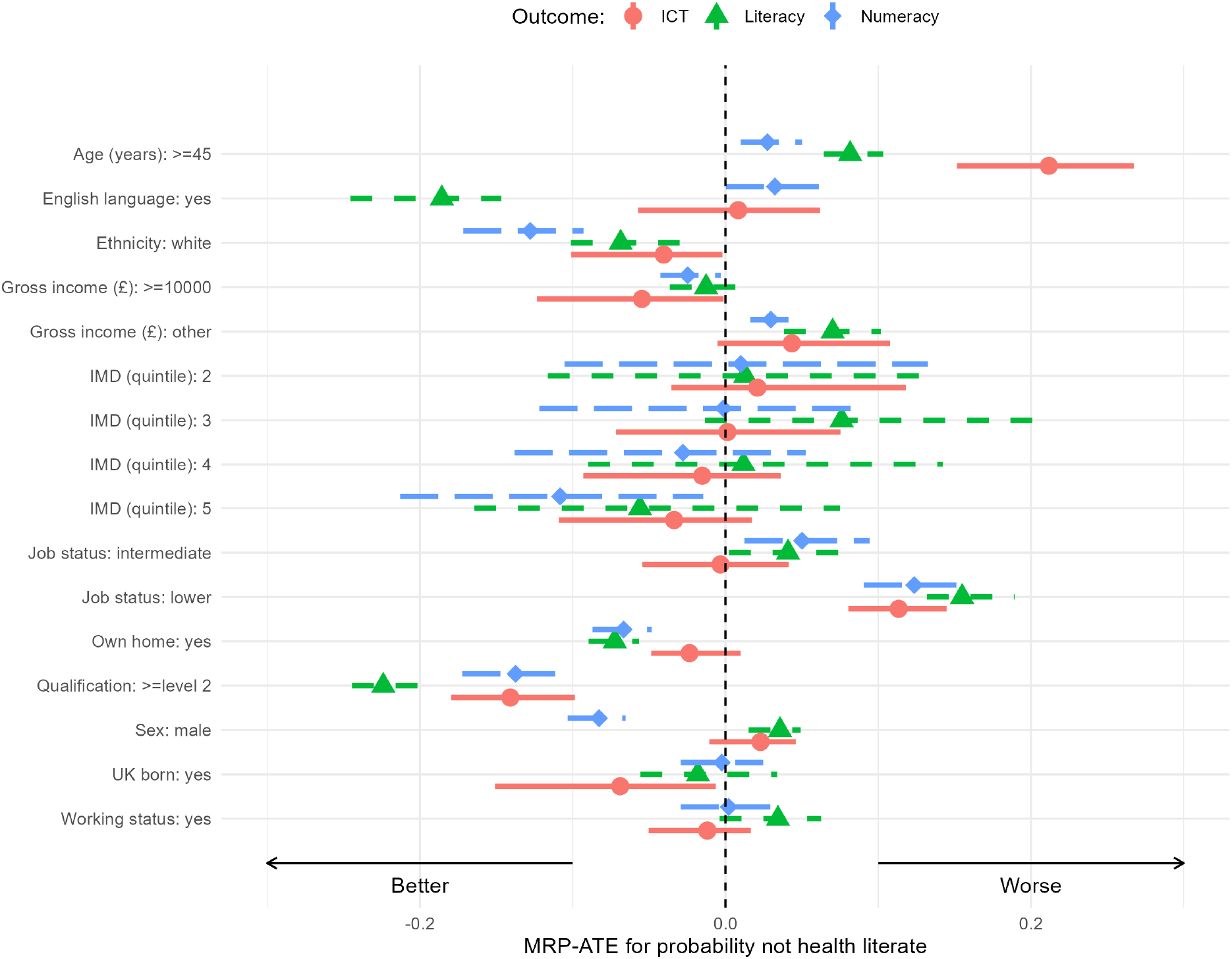
Forest plot of MRP-ATE for probability of not health literate for each of the outcomes ICT (red circles), literacy (green triangles) and numeracy (blue squares).

The plot shows that the ICT outcome is not impacted as significantly as literacy and numeracy when changes in covariate values occur. The exception to this is age where people over 45 years old have a markedly worse change in ICT scores compared to literacy and numeracy scores (0.192 [0.146, 0.258]). Numeracy scores are impacted most by a change to White ethnicity and qualification level being level 2 or higher (−0.129 [−0.160, −0.095] and −0.141 [−0.166, −0.117] respectively). All outcomes (ICT, literacy, and numeracy) were impacted positively by having a qualification at level 2 or above − the only factor to have a wholly positive impact across all three outcomes. The ICT outcome tends to have smaller uncertainty for IMD and no improvement for less deprived areas compared to the other outcomes. For literacy and numeracy, less deprived areas indicate improvement but are clearly not significant. ICT and in particular literacy have worse health literacy estimates for more deprived quintiles relative to quintile 1 (most deprived). The IMD results exhibit borrowed strength and shrinkage to the pooled mean due to the Bayesian multilevel property which regularised the fit. Recall, there are few Newham residents in quintile 1 thus pulling it more towards the pooled mean value and most of the residents are in quintiles 2 and 3. Counter-intuitively, quintile 3 for literacy appears to actually be worse than quaitile 1; however, a model with fixed effect for IMD gives an OR of 0.74 indicating a consistent improvement for less deprived areas, which corresponds with Rowlands et al (2015). Male sex shows an increase in numeracy (−0.092 [−0.119, −0.066]). Unsurprisingly, English as a first language has a strong benefit for literacy (−0.171 [−0.217, −0.130]) but little to no influence on the other health literacy outcomes.

Figure 3 shows rankogram plots of cumulative ranks probabilities *P*_*ijr*_. These plots show the likelihood of a variable being ranked 1st to 4th as the variable which will have the most impact on the outcome (ICT, literacy, or numeracy) in terms of change in MRP-ATE. This allows us to make direct comparison between the different covariates accounting for both the effect size and uncertainty. We restrict the plot to only show the variables with a minimum cumulative rank of at least 0.25 so that the plot is clearer and focuses on the most important variables. Recall that the absolute treatment effects are used to rank so the magnitude but not direction of the MRP-ATE relative to reference category is indicated. The full rankogram plot with all categories and all ranks is given in the Appendix Figure A.4.

**Fig. 3.**
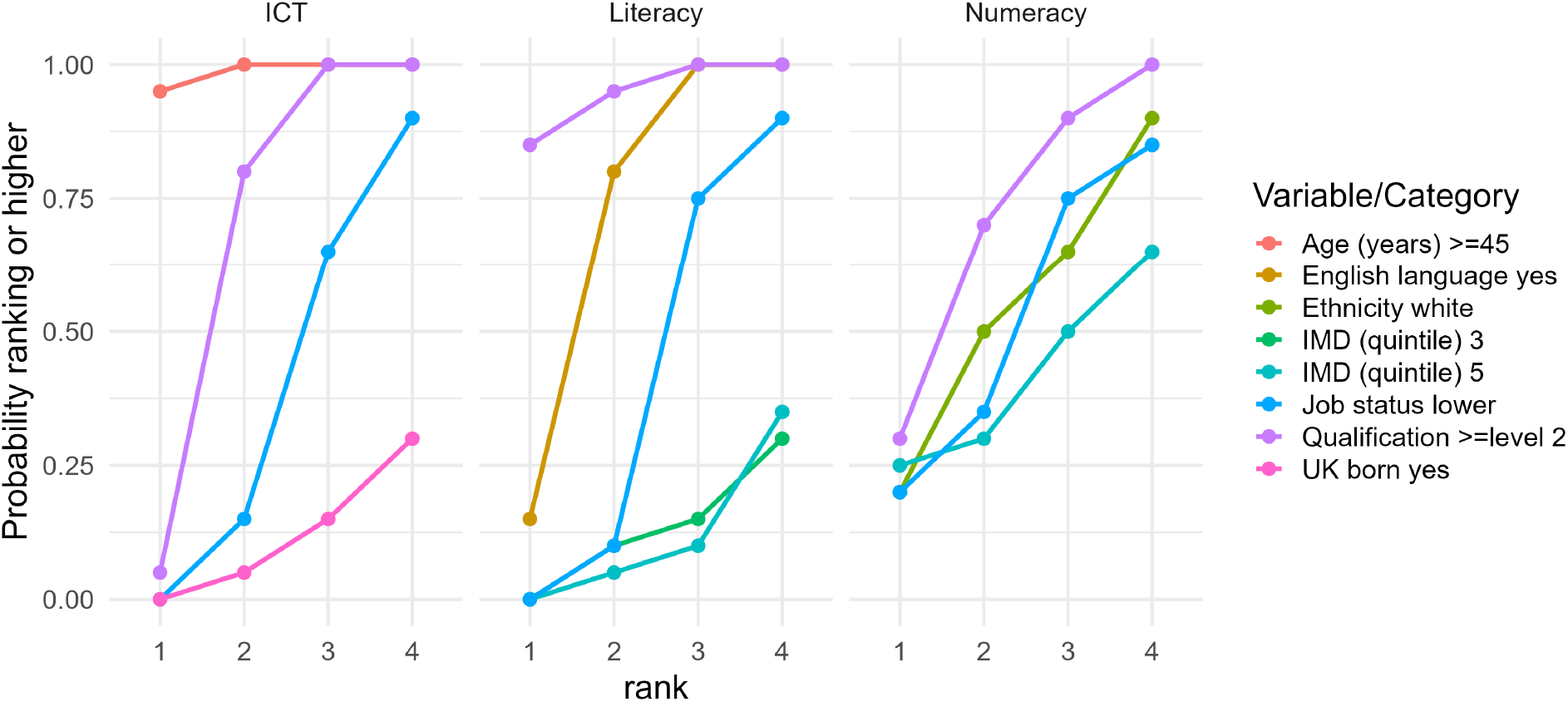
Rankogram plots of cumulative rank probabilities for the first four positions and each of the outcomes of not health literate for ICT, literacy and numeracy. Only probabilities above 0.25 are shown for clarity. MRP-ATE for absolute treatment effects are used to rank so magnitude and not direction of effect relative to reference category is indicated.

**Fig. 4.**
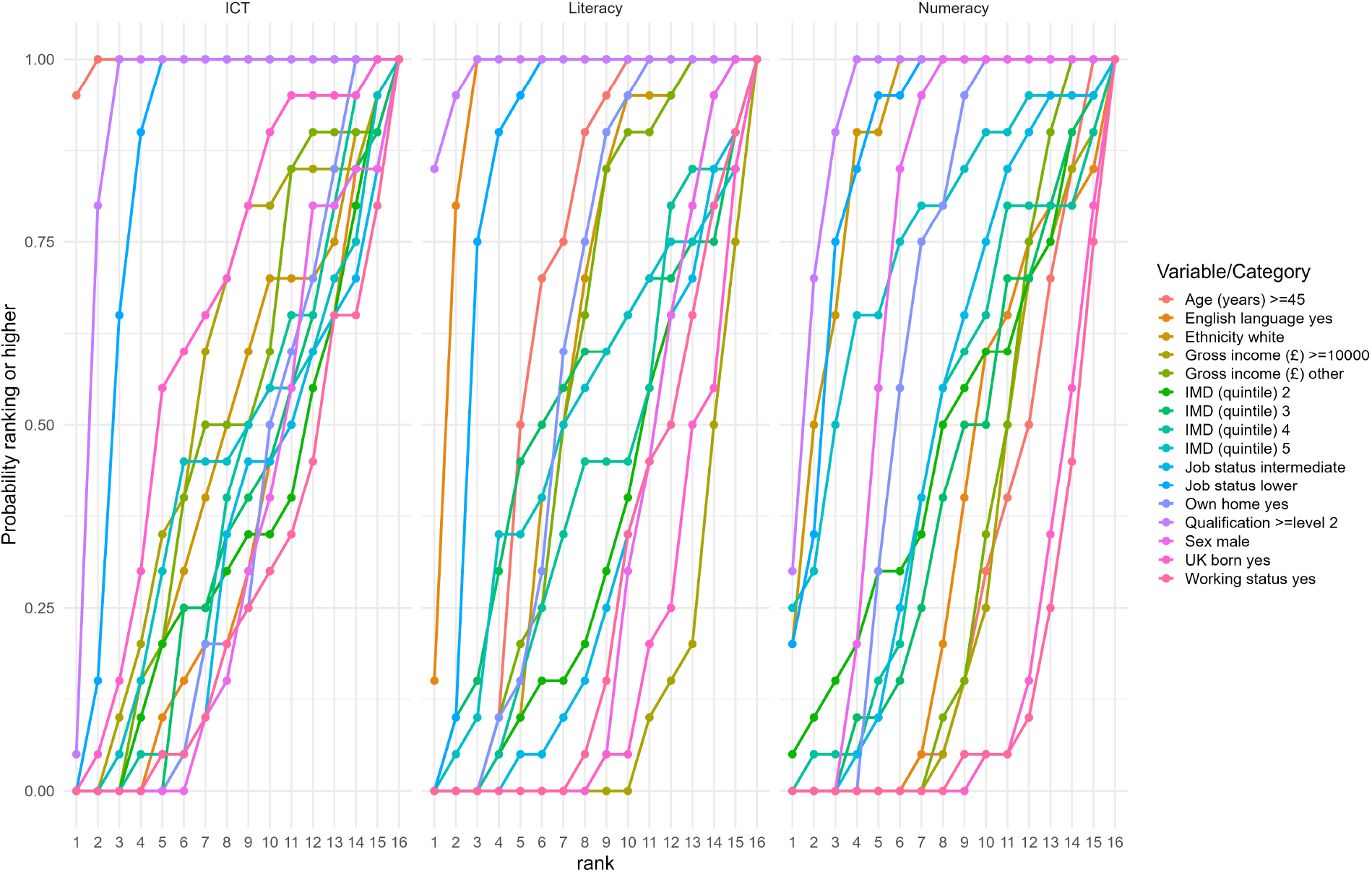
Cumulative rank probability plots for absolute MRP-ATE outcomes for all ranks and all variable categories.

For ICT, the *≥*45 age threshold clearly has the highest chance of ranking in the top positions. The next two best ranking variables are clearly qualification then lower job status. For literacy, qualification has the highest probability of ranking first. English as first language has the next highest probability of ranking second. Finally, for numeracy, it is less clear cut which variable is the top ranking. Qualification is the top ranking but with less certainty with both white ethnicity and lower job status also competing. A lack of deprivation, i.e. upper IMD quintile, is also not insignificant.

Similar to the rankogram plot, Table 3 shows the SUCRA and expected rank statistics for each variable and health literacy outcome using the absolute value of the MRP-ATE estimates. The values correspond between the two statistics since they are both derived from the cumulative rank. Qualification greater than level 2 ranks first or second across all outcomes. English language ranks second for literacy, and age ≥45 years old ranks first for ICT.

**Table 3.** SUCRA and expected rank using absolute value MRP-ATE for the health literacy outcomes ICT, literacy and numeracy.

| Variable | Category | SUCRA | | | $\mathbb{E}[\text{rank}]$ | | |
| --- | --- | --- | --- | --- | --- | --- | --- |
|  |  | ICT | Literacy | Numeracy | ICT | Literacy | Numeracy |
| Age (years) | $\geq 45$ | 100 | 66 | 27 | 1 | 6 | 12 |
| English Language | Yes | 33 | 93 | 34 | 11 | 2 | 11 |
| Ethnicity | White | 46 | 56 | 88 | 9 | 8 | 3 |
| Gross Income (£) | $\geq 10000$ | 55 | 11 | 28 | 8 | 14 | 12 |
|  | Other | 49 | 55 | 32 | 9 | 8 | 11 |
| IMD (quintile) | 2 | 34 | 35 | 47 | 11 | 11 | 9 |
|  | 3 | 36 | 51 | 40 | 11 | 8 | 10 |
|  | 4 | 39 | 40 | 45 | 10 | 10 | 9 |
|  | 5 | 43 | 49 | 74 | 10 | 9 | 5 |
| Job Status | Intermediate | 31 | 30 | 49 | 11 | 12 | 9 |
|  | Lower | 85 | 85 | 87 | 3 | 3 | 3 |
| Own Home | Yes | 36 | 58 | 62 | 11 | 7 | 7 |
| Qualification | ≥Level 2 | 92 | 99 | 93 | 2 | 1 | 2 |
| Sex | Male | 32 | 28 | 70 | 11 | 12 | 5 |
| UK Born | Yes | 63 | 16 | 13 | 6 | 14 | 14 |
| Working Status | Yes | 26 | 26 | 11 | 12 | 12 | 14 |

## Discussion

We estimated an increase in numeracy health literacy for UK born. This may be due to non-UK born may have received their primary and secondary education in different countries with varying curricula and teaching standards, which could affect their performance on a UK-based skills assessment. Broader societal issues, such as systemic biases within the education system or other institutions, can create disadvantages for ethnic minority groups. These disadvantages can manifest as disparities in access to high-quality education, resources, and support, which in turn affect the development of skills such as numeracy.

The health materials used to set the competency thresholds were assessed for both their complexity in literacy and numeracy. Numeracy tests, especially those based on real-world scenarios such as health information, are not just about pure calculation. They often involve word problems that require strong reading comprehension to understand the context and what is being asked. Therefore, lower English literacy skills could directly hinder performance on a numeracy test, even if the person’s mathematical ability is strong. Finally, it may be that societal factors and stereotypes can sometimes create higher levels of “maths anxiety” in certain demographic groups Maloney and Beilock (2012). This can affect confidence and performance in testing situations specifically related to mathematics and numeracy, independent of other academic skills.

The decrease in health literacy for literacy may be a spurious result for this data set or it may indicate something about a focus of government funding, non-profit aid, and specific support programs on the most deprived areas and the next most deprived may be falling between the cracks.

We also recognise that the ethnicity categorisation as taken from the SfL survey is overly simplistic (originally coded White or Black and minority ethnic (BME) only). This is an increasingly outdated approach that masks significant diversity and is a significant limitation for diverse populations in areas such as Newham.

We showed that a more deprived area tends to correlate to a decrease in health literacy outcomes. This corresponds with other more specific variables since IMD can be thought of as a compound measure. IMD includes a domain for education, skills, and training deprivation. Less deprived areas scores often have lower levels of educational attainment. We overwhelmingly identify qualification level as the most influential factor across all health literacy outcomes. Therefore, the educational disadvantage captured by IMD directly translates to lower literacy and numeracy skills, which are the core components of the health literacy definition used in this research. Limited access to resources includes less access to computers and the internet; reduced ability to afford stable housing, adequate or high quality nutrition, and other resources that support overall well-being as well as the capacity to engage with health services potentially due to limited time and energy to focus on preventive health or decipher complex health information due to the stresses of financial instability and day-to-day challenges and pressures. It has also been recognised that people with more social complexity and inequity driven by systemic inequality may also have reduced levels of trust in institutions and systems as they have failed to support them previously, further affecting access and capacity to engage with health services NHS Race and Health Observatory (2024).

The University of Southampton and NHS England created a web tool to estimate the percentage of a local authority population with low health literacy and numeracy. It also used the 2011 Skills for Life Survey with a population update based on the 2021 Census and the 2019 Index of Deprivation University of Southampton and NHS England (n.d.). Their estimated health literacy prevalence in Newham was 74.78% (second lowest in England). Newham’s neighbouring boroughs included Barking and Dagenham 78.41%, Tower Hamlets 67.61% and Greenwich 66.35%. The national average was 58.30%. These boroughs are somewhat similar to Newham and its population’s demographics, with Tower Hamlets being the most comparable.

## Conclusions

This study provides an extensive, qualitative analysis of the determinants of health literacy in Newham, London, using advanced statistical techniques and multiple data sets. The research identifies several key factors that significantly impact health literacy outcomes, with implications for targeted interventions and policy-making in the area.

Health literacy in this study is defined using the Skills for Life (SfL) 2011 survey, encompassing literacy, numeracy, and ICT skills. This definition allows for a comprehensive assessment of health literacy, moving beyond traditional unidimensional measures. The analysis combines data from the SfL and LFS surveys, the Newham Residents Survey (NRS), and the UK 2021 Census. The NRS and Census data provide local demographic information. This combination of data sets addresses the limitations of each individual source, allowing a more complete understanding of the factors that influence health literacy in Newham.

We employed multilevel regression with post-stratification (MRP) to estimate health literacy at the subnational level, to provide more precise estimates for small areas and subgroups. The research uses average treatment effects (ATE) to quantify the impact of various factors on health literacy, providing insights into how changes in these variables affect health literacy outcomes. Key determinants of health literacy were identified by Rowlands et al (2015), including age, ethnicity, qualification level, English as a first language, job status, gross income, and home ownership. These factors have varying impacts on different aspects of health literacy.

A motivation for ranking the determinants of health literacy was to facilitate communication with public health professionals. By translating complex statistical outputs into a more intuitive format, ranking helps to clearly identify which socio-demographic factors have the most substantial impact on literacy, numeracy, and ICT skills within Newham. This prioritisation is crucial for effectively allocating limited resources. However, ranking does simplify multifaceted issues and the relative importance of factors can be sensitive to the specific statistical model and data used. Therefore, rankings can be a powerful communication tool, but are only a guide.

Health literacy interventions at the individual level include, educational programs, simplified health communication strategies, digital health tools, and numeracy training to improve comprehension and decision-making. Healthcare system-level interventions include training providers in clear communication, screening for low health literacy, offering patient navigation services, and ensuring culturally and linguistically appropriate materials. Community-based and policy interventions involve integrating health literacy into schools and workplaces, public health campaigns, partnerships with local organizations, and advocating for policies that promote equitable access to health information. These strategies aim to empower individuals and improve health outcomes.

With new plans for the NHS being published recently, the need to pivot the English health system to a prevention-first approach, improving health literacy of institutions and individuals is a key opportunity and underpinning factor that will need to be addressed to improve access, knowledge, and experience for patients, staff, and the public. This data and any future tool created will help the stakeholders in a local area to understand the factors that most impact health literacy, and therefore enable them to consider how they can adjust their practice or deliver an intervention which can improve health literacy for the residents they work with. At a time when local authority budgets and resources are stretched and limited or being reduced, being able to focus the limited resources on interventions more likely to have an impact supports best practice for effective and efficient use of public funds. The results of this work can inform targeted health literacy interventions in Newham, focussing on the groups and areas with the greatest impact. An up-to-date quantification can better guide resource allocation and policy decisions, ensuring that interventions are tailored to the specific needs of the community.

There are several key directions for future research and practice.

Building on previous work, such as the Geodata app University of Southampton and NHS England (n.d.), future work should look to develop an accessible, interactive web tool with the methods presented here, such as a Shiny app, to allow stakeholders to explore their own data and context dynamically, enabling a more detailed, evidence-based approach to developing new health literacy interventions.

Alternative or additional existing data sets, such as those available from the ONS Understanding Society UK Household Longitudinal Study or PIAAC may be considered in the future. Further, as local authorities increase the coproduction and data collection they undertake both internally and with residents, there are opportunities to collect additional local data which will provide more locally accurate and up-to-date data, e.g., the Newham Residents Survey. This could be used to validate the methods and extended to provide the full covariate correlation structure required for MRP. To address not currently have access to this, we employed iterative proportional fitting (IPF) in order to synthetically simulate the required data using marginal summary statistics. Alternative methods are available and this is an area of active research Pitts et al (2025).

It will be necessary to align with best practice work which is developed nationally, regionally, and locally to ensure the model iterates to incorporate the data which can evidence the impact of best practice locally. Therefore, it would be beneficial to consider measures which show change quickly (e.g. annual data collection) that could be included in the model to enable close to real-time monitoring of interventions to evidence impact and inform iterations.

In addition, more granular data (e.g. ethnicity) and more culturally appropriate or sensitive data collection methods will provide additional insights to inform community-specific approaches and provide a more accurate understanding of how standardised measurements such as the SfL may affect the measures of ICT, literacy, and numeracy (e.g. English comprehension being required to complete mathematics questions). This will also help institutions to assess where different approaches may be necessary for different groups of patients.

The ATE estimates as used in our analysis provided the largest change on the outcome scale between two discrete category counterfactual scenarios. This is natural in terms of a rate of change or gradient, as in an average marginal effect model. For our context, an alternative approach could be to use the *average treatment effect on treated (ATT)*. This is the ATE for only the subset of population that would have their covariate value changed and does not include those who already have that value. Further, there exist ATE variants for heterogenous, marginal and quantile treatment effects. Finally, additional modifications can be made to account for relative population sizes, importance or budgets.

We also assumed that it is possible to intervene on a covariate without affecting the remainder. It may be that a direct change to one covariate has an effect on the other covariates which in turn has an indirect effect on the outcome Díaz and Hejazi (2020). It may also be that possible values available for an intervention for an individual may depend on their particular set of covariates. Finally, we have assumed deterministic interventions but this may be generalised to *stochastic* interventions, loosely defined as interventions that yield a random variable focal covariate.

Up-to-date, local, detailed data should be collected to best inform specific questions. In turn, all available evidence should be principally synthesised if relevant even indirectly; a problem suited to a Bayesian paradigm. For example, the current analysis could be extended to include multiple surveys over time or location to capture temporal and spatial variation and allow future predictions. Further, research is needed to quantify the cost-effectiveness of health literacy initiatives and to rigorously evaluate their long-term impact on clinical and health outcomes.

There is an ongoing need for more sensitive and comprehensive measures of health literacy, particularly for diverse populations such as Newham and within evolving digital health environments. This includes developing methods to accurately assess digital health literacy among digitally marginalized groups. For example, development of innovative tools, such as AI-driven text simplification or translation, could make health information more accessible. However, rigorous evaluation is necessary to ensure accuracy and comprehensibility of such technologies.

Health literacy, and any interventions aim at addressing it, are closely linked to health patient activation. This refers to an individual’s confidence and motivation to manage their own health and healthcare. The Patient Activation Measure (PAM) is a common validated tool that assesses this, ranging from passive and overwhelmed to proactive and confident. It can be used as a basis to develop interventions targeted at communities most of risk of missing out from likely gains in digital health such as the NHS app.

Our work will help inform targeted and tailored interventions to prioritize the design and implementation of community-based, culturally appropriate interventions. Incorporating health literacy screening into standard healthcare practices can help identify patients needing additional support and facilitate timely provider responses. However, it should be noted that the potential for driving inequalities and inequities will need to be mitigated against (e.g. preventing biases and stigma due screening outcomes). Furthermore, in populations with low trust in the ‘system’ much of the data used in the model may not be available on every patient record which would limit the accuracy and effectiveness of screening tools that rely on demographic data and the like, especially those not regularly collected or updated (e.g., employment) in health and care records.

Alongside individual health literacy development, there must be a sustained focus on improving *organizational* health literacy. This involves ensuring that healthcare systems and public health organisations are designed to provide clear, accessible and culturally and linguistically appropriate health information and services in all communications (including face-to-face or verbal sharing of information). As new determinants of health emerge and become more understood, such as digital and commercial determinants, continued research and policy attention are required to understand their impact on health literacy and to develop proactive strategies to mitigate potential inequities.

## Data Availability

All data produced are available online at https://github.com/n8thangreen/healthliteracy

## Funding

Not applicable

## Ethics approval and consent to participate

This study was a secondary analysis of anonymized data from the Newham Residents Survey, provided by the London Borough of Newham. The original survey and its data collection protocols were managed by the London Borough of Newham, which was responsible for obtaining informed consent from all participants at the time of data collection. The survey was compliant with the London Borough of Newham’s information governance and Data Protection, and GDPR rules and regulations. The survey was conducted by Opinion Research Services (ORS). All procedures were carried out in accordance with the ethical guidelines of the Market Research Society (MRS) and the company’s internal research governance and quality assurance procedures. All other data were obtained from openly-available ONS resources. In accordance with the research governance policies of UCL, research using non-identifiable, pre-existing data is exempt from formal ethics review. Therefore, a separate ethics approval was not required for this study. All methods were carried out in accordance with relevant guidelines and regulations, including the UK Data Protection Act 2018.

## Appendix

## Notes

### Competing Interest Statement

The authors have declared no competing interest.

### Funding Statement

This study did not receive any funding

### Author Declarations

London Borough of Newham gave ethical approval for this work. Opinion Research Services (ORS) gave ethical approval for this work. This study was a secondary analysis of anonymized data from the Newham Residents Survey, provided by the London Borough of Newham.

